# A Comprehensive Analysis of COVID-19 Transmission and Fatality Rates at the County level in the United States considering Socio-Demographics, Health Indicators, Mobility Trends and Health Care Infrastructure Attributes

**DOI:** 10.1101/2020.08.03.20164137

**Authors:** Tanmoy Bhowmik, Sudipta Dey Tirtha, Naveen Chandra Iraganaboina, Naveen Eluru

**Affiliations:** Department of Civil, Environmental & Construction Engineering, University of Central Florida

**Keywords:** COVID-19, transmission rate, mortality rate, linear mixed model, policy analysis, vulnerable counties

## Abstract

**Background:** Several research efforts have evaluated the impact of various factors including a) socio-demographics, (b) health indicators, (c) mobility trends, and (d) health care infrastructure attributes on COVID-19 transmission and mortality rate. However, earlier research focused only on a subset of variable groups (predominantly one or two) that can contribute to the COVID-19 transmission/mortality rate. The current study effort is designed to remedy this by analyzing COVID-19 transmission/mortality rates considering a comprehensive set of factors in a unified framework.

**Method:** We study two per capita dependent variables: (1) daily COVID-19 transmission rates and (2) total COVID-19 mortality rates. The first variable is modeled using a linear mixed model while the later dimension is analyzed using a linear regression approach. The model results are augmented with a sensitivity analysis to predict the impact of mobility restrictions at a county level.

**Findings:** Several county level factors including proportion of African-Americans, income inequality, health indicators associated with Asthma, Cancer, HIV and heart disease, percentage of stay at home individuals, testing infrastructure and Intensive Care Unit capacity impact transmission and/or mortality rates. From the policy analysis, we find that enforcing a stay at home order that can ensure a 50% stay at home rate can result in a potential reduction of about 30% in daily cases.

**Interpretation:** The model framework developed can be employed by government agencies to evaluate the influence of reduced mobility on transmission rates at a county level while accommodating for various county specific factors. Based on our policy analysis, the study findings support a county level stay at home order for regions currently experiencing a surge in transmission. The model framework can also be employed to identify vulnerable counties that need to be prioritized based on health indicators for current support and/or preferential vaccination plans (when available).

**Funding:** None.

**RESEARCH IN CONTEXT:** *Evidence before this study:* We conducted an exhaustive review of studies examining the factors affecting COVID-19 transmission and mortality rates at an aggregate spatial location such as national, regional, state, county, city and zip code levels. The review considered articles published in peer-reviewed journals (via PubMed and Web of Science) and working articles uploaded in preprint platforms (such as medRxiv). A majority of these studies focused on a small number of counties (up to 100 counties) and considered COVID-19 data only up to the month of April. While these studies are informative, cases in the US grew substantially in recent months. Further, earlier studies have considered factors selectively from the four variable groups - <u>socio-demographics, health indicators, mobility trends</u>, and <u>health care infrastructure attributes. The exclusion of variables from these groups</u> is likely to yield incorrect/biased estimates for the factors considered.

*Added value of this study:* The proposed study enhances the coverage of COVID-19 data in our analysis. <u>Spatially</u>, we consider 1258 counties encompassing 87% of the total population and 96% of the total confirmed COVID-19 cases. <u>Temporally</u>, we consider data from March 25^th^ to July 3^rd^, 2020. The model system developed comprehensively examines factors affecting COVID-19 from all four categories of variables described above. The county level daily transmission data has multiple observations for each county. To accommodate for these repeated measures, we employ a linear mixed modeling framework for model estimation. The model estimation results are augmented with policy scenarios imposing hypothetical mobility restrictions.

*Implications of all the available evidence:* The proposed framework and the results can allow policy makers to (a) evaluate the influence of population behavior factors such as mobility trends on virus transmission (while accounting for other county level factors), (b) identify priority locations for health infrastructure support as the pandemic evolves, and (c) prioritize vulnerable counties across the country for vaccination (when available).

## INTRODUCTION

Coronavirus disease 2019 (COVID-19) pandemic, as of July 19^th^, has spread to 188 countries with a reported 14.4 million cases and 603 thousand fatalities^1^. The pandemic has affected the mental and physical health of people across the world significantly taxing the social, health and economic systems^2^. Among the various countries affected, United States has reported the highest number of confirmed cases (3.8 million) and deaths (140 thousand) in the world^3^. In this context, it is important that we clearly understand the factors affecting COVID-19 transmission and mortality rate to prescribe policy actions grounded in empirical evidence to slow the spread of the transmission and/or prepare action plans for potential vaccination programs in the near future. Towards contributing to these objectives, the current study develops a comprehensive framework for examining COVID-19 transmission and fatality rates in the United States using COVID-19 data at a county level encompassing about 87% of the US population. The study effort is designed with the objective of including a universal set of factors affecting COVID-19 in the analysis of transmission and mortality rates. We employ an exhaustive set of county level characteristics including (a) <u>socio-demographics</u>, (b) <u>health indicators</u>, (c) <u>mobility trends</u>, and (d) <u>health care infrastructure attributes</u>. We recognize that analysis of COVID-19 data with a subset of factors, as has been the case with earlier work, is likely to yield incorrect/biased estimates for the factors considered. The framework proposed for understanding and quantifying the influence of these factors can allow policy makers to (a) evaluate the influence of population behavior factors such as mobility trends on virus transmission (while accounting for other county level factors), (b) identify priority locations for health infrastructure support as the pandemic evolves, and (c) prioritize vulnerable counties across the country for vaccination (when available).

In recent months, a number of research efforts have examined COVID-19 data in several countries to identify the factors influencing COVID-19 transmission and mortality. Given the focus of our current study, we restrict our review to studies that explore COVID-19 transmission and mortality rate at an aggregated spatial scale. To elaborate, these studies explored COVID-19 transmission and mortality rates at the national^4–6^, regional^7^, state^8^, county^9–14^, city^15^ and zip code levels^16^. A majority of these studies considered transmission rate as the response variable (transmission rate per capita). The main approach employed to identify the factors affecting the response variables is the linear regression approach. In their analysis, researchers employed a host of independent variables from four variable categories: socio-demographics, health indicators, mobility trends and health care infrastructure attributes. For socio-demographics, studies found income, race and age distribution have a positive association with the COVID-19 transmission ^11,16–18^. Regarding health indicators, earlier research found that smokers, obese and individuals with existing health conditions are more likely to be severely affected by COVID-19^11^. In terms of mobility trends, studies showed that staying at home and effective mobility restriction measures significantly lower the COVID-19 transmission rate^4,7,9,10,14^ while increased mobility resulted in increased COVID-19 transmission^12,19^. Finally, among health care infrastructure attributes, testing rate is linked with reduced risk of COVID-19 transmission^4,5^. While earlier research efforts have considered the factors from all variable categories, it is important to recognize that each individual study focused only on a subset of variable groups (predominantly one or two) and have not controlled explicitly for other variable groups that can contribute to the COVID-19 transmission/mortality rate.

The current study builds on earlier literature examining the factors affecting COVID-19 transmission and mortality rate and contributes along the following directions. *First*, we extensively enhance the spatial and temporal coverage of COVID-19 data in our analysis. *Spatially*, earlier research on COVID-19 aggregate data has focused on a small number of counties (up to 100 counties). In our study, we consider all counties with total number of cases greater than 100 on July 3^rd^. The 1258 counties selected encompass 87% of the total population and 96% of the total confirmed COVID-19 cases. *Temporally*, earlier research has only considered data up to the month of April. While these studies are informative, cases in the US grew substantially in the recent months. Hence, in our study we have considered data from March 25^th^ to July 3^rd^, 2020. The longer period of data (101 days) also enables us to study/test for the evolution of variable effects over time. *<u>Second</u>*, earlier research studies have considered factors from one or two of the categories of variables identified above. Further, studies that tested health indicators employed one or two measures selectively. In our analysis, we conduct a comprehensive examination of factors affecting COVID-19 from all four categories of variables including (a) <u>socio-demographics</u>: distribution by age, gender, race, income, location (urban or rural), education status, income inequality and employment, (b) <u>health indicators</u>: percentage of population suffering from cancer, cardiovascular disease, hepatitis, Chronic Obstructive Pulmonary Disease (COPD); diabetes, obesity, Human Immunodeficiency Virus (HIV), heart disease, kidney disease, asthma; drinking and smoking habits, (c) <u>mobility trends</u>: daily average exposure, social distancing matrices, percentage of people staying at home, and (d) <u>health care infrastructure attributes</u>: hospitals per capita, ICU beds per capita, COVID-19 testing measures. *<u>Finally</u>*, the research study employs a robust modeling framework in terms of model structure and dependent variable representation. A linear mixed model system that addresses the limitations of the traditional linear regression framework for handling repeated measures is employed. For dependent variable, alternative functional forms of COVID-19 transmission – natural logarithm of daily cases per 100 thousand people and natural logarithm of 7-day moving average of cases per 100 thousand people - are considered in model estimation. The overall approach allows us to robustly quantify the impact of factors affecting COVID-19 transmission.

## METHODS

### Data Collection

#### Independent variables

Table 1 summarizes sample characteristics of the explanatory variables with the definition considered for final model estimation, the data source, and sample characteristics (minimum, maximum and mean values). For the sake of brevity, in the ensuing discussion we only present details of two groups of independent variables: health indicators and mobility trends. Using health indicator data, we ranked the 1,258 counties in a descending order of health metric and provided it in Figure 1. Further, we compute the average values for different health indicators across the healthiest and unhealthiest 10 counties to highlight the change in health conditions across the two groups. The values clearly emphasize the vulnerability of the unhealthiest counties relative to the healthiest counties. For instance, number of HIV patients in the healthy counties are 75·83 while in the unhealthiest counties, it is almost 430% higher (407).

**Table 1.** Descriptive Statistics of the Dependent and Independent Variables.

| Variables | Source | Mean | Min/Max | Sample Size |
| --- | --- | --- | --- | --- |
| **Independent Variables** |  |  |  |  |
| **Demographic Characteristics** |  |  |  |  |
| Percentage of population aged 18 years and lower | ACS<sup>a</sup> | 22.705 | 7.155/33.882 | 1258 |
| Percentage of population aged 65 years and over | ACS | 16.644 | 7.545/56.944 | 1258 |
| Percentage of African American | ACS | 13.133 | 0.113/80.507 | 1258 |
| Percentage of Hispanic | ACS | 11.331 | 0.623/96.323 | 1258 |
| Percentage of Female | ACS | 50.507 | 37.041/54.495 | 1258 |
| Ln (Median income) | ACS | 10.903 | 10.150/11.820 | 1258 |
| Percentage of people less than high school education | ACS | 14.003 | 3.127/47.053 | 1258 |
| Employment rate per capita | ACS | 0.446 | 0.195/0.634 | 1258 |
| Income inequality ratio (80<sup>th</sup> percentile/20<sup>th</sup> percentile) | CHRR<sup>b</sup> | 4.574 | 3.150/9.148 | 1258 |
| **Health Indicators** |  |  |  |  |
| Ln (HIV Prevalence Rate per 100K people) | CHRR | 5.043 | 0.723/7.859 | 1258 |
| Hepatitis B Cases per 100K people in2017 | CDC<sup>c</sup> | 1.250 | 0.000/11.700 | 1258 |
| Hepatitis C Cases per 100K people in2017 | CDC | 0.980 | 0.000/5.600 | 1258 |
| Asthma % for >= 18 years | CDC | 9.328 | 7.400/12.300 | 1258 |
| COPD % for >= 18 years | CDC | 6.692 | 3.300/13.700 | 1258 |
| Reported cancer case per 100K people | CDC | 455.918 | 241.000/576.400 | 1258 |
| Percentage of diabetic | CHRR | 11.353 | 3.300/20.400 | 1258 |
| Percentage of obesity among adults | CHRR | 31.653 | 13.600/46.700 | 1258 |
| Cardiovascular Disease Hospitalization Rate per 1,000 Medicare Beneficiaries | CDC | 63.609 | 0.300/110.500 | 1258 |
| **Mobility Trends** |  |  |  |  |
| Ln (Daily Average Exposure), 10 days lag |  |  |  |  |
| From April 25th | CEI<sup>d</sup> | 3.325 | 0.102/0.644 | 127058 |
| % People staying at home |  |  |  |  |
| 14 days lag | Safegraph | 4.128 | 0.847/7.049 | 127058 |
| **Healthcare Related Attributes** |  |  |  |  |
| Hospitals per 100K people | CHRR | 1.967 | 0.000/15.644 | 1258 |
| Number of ICU beds per capita | CHRR | 20.111 | 0.000/171.850 | 1258 |
| Ln (No of tests with 5 days lag) | CTP<sup>e</sup> | 11.178 | 4.320/15.190 | 5151 |
| **Temporal Factors** |  |  |  |  |
| Day is weekend | -- | 0.277 | 0.000/1.000 | 127058 |
| **Dependent Variables** |  |  |  |  |
| Ln (Daily COVID-19 transmission rate per 100K people) | CSSE<sup>f</sup> | 1.455 | 0.000/7.670 | 127058 |
| Ln (Total COVID-19 mortality rate per 100K people) | CSSE | 2.722 | 0.000/7.120 | 1258 |

<sup>a</sup> = American Community Survey<sup>b</sup> = County Health Rankings & Roadmaps<sup>c</sup> = Central for Disease Control System<sup>d</sup> = COVID Exposure Indices<sup>20</sup><sup>e</sup> = COVID-19 Tracking Project<sup>29</sup><sup>f</sup> = Center for Systems Science and Engineering Coronavirus Resource Center at Johns Hopkins University<sup>21</sup>

**Figure 1.**
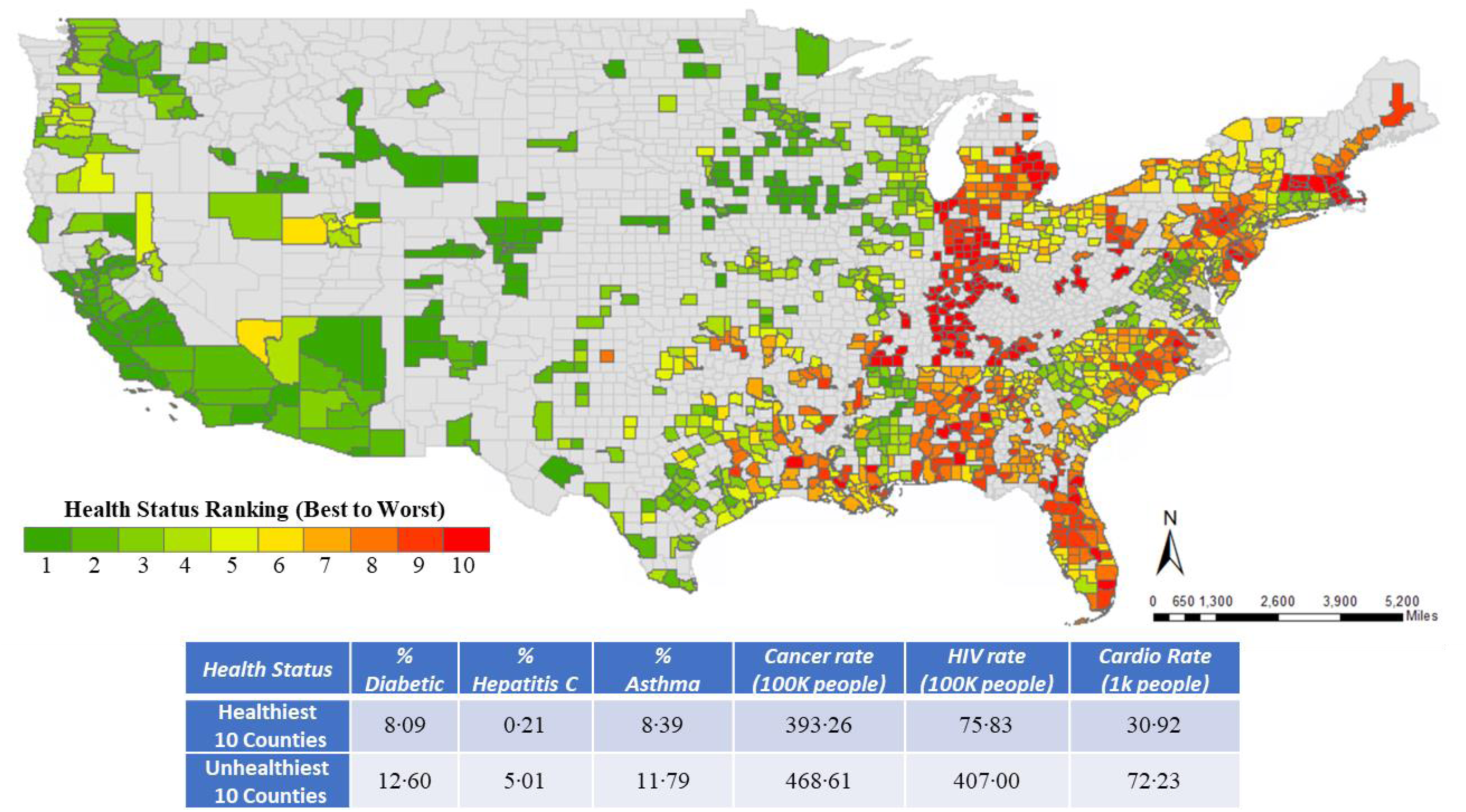
Ranking of Counties Based on Health Indicators.

To incorporate mobility trends, we considered two variables: daily average exposure^20^ and social distancing metric (from SafeGraph^1^) to serve as surrogate measures for the mobility patterns (see Supplementary Material for detail). Figure 2 provides a summary of both these measures at a state level from January 22^nd^ to July 3^rd^. From the figure, we can clearly see the reduction in average daily exposure in March as many states and local jurisdictions imposed lockdowns. By late April, exposure activity started to increase again across all the states while still being lower than the levels for February. In terms of the staying at home measure, as expected, we find an exactly opposite trend.

**Figure 2.**
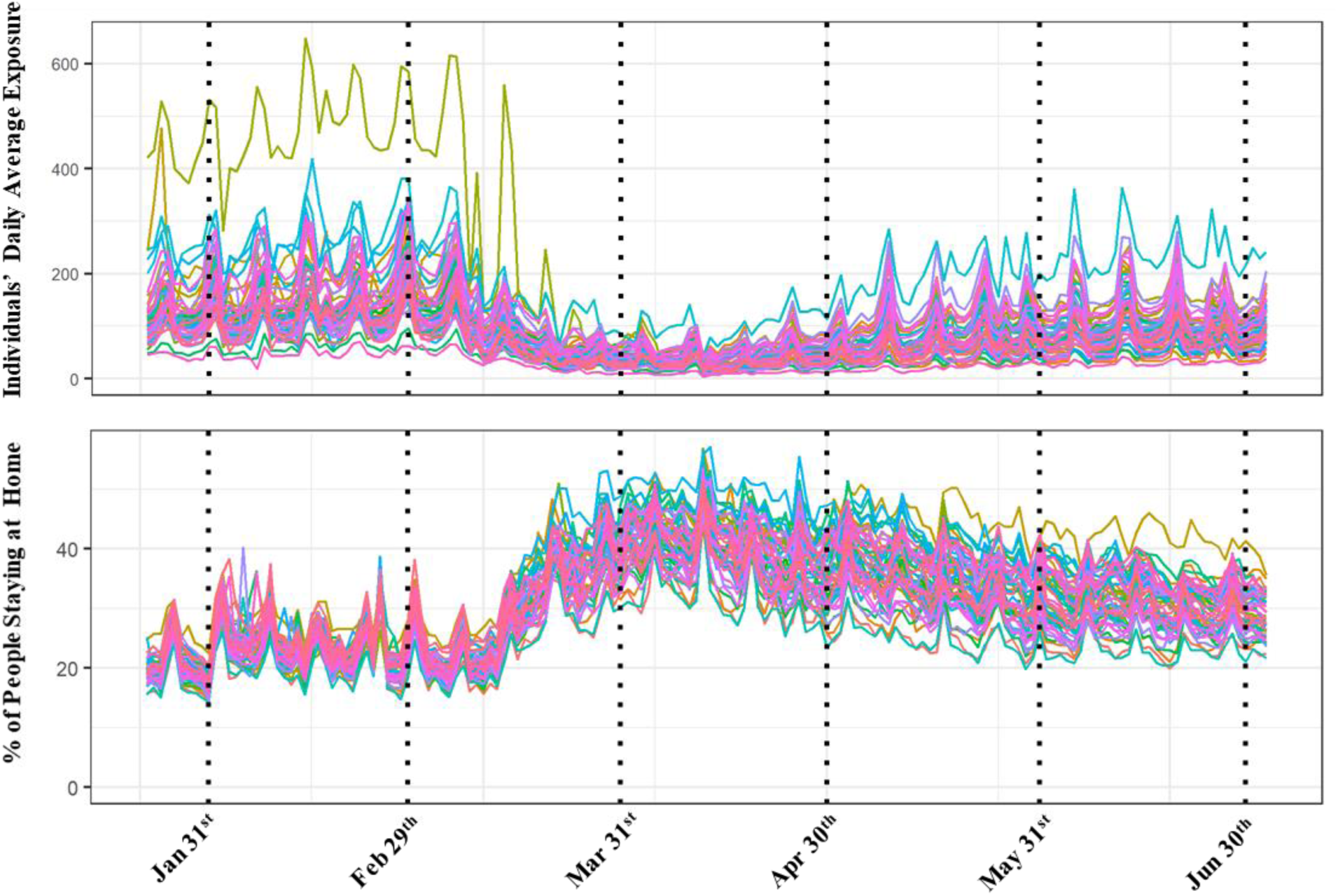
Average Daily Exposure and Percentage of People Staying at Home.

#### Dependent variables

We analyze two county level dependent variables: (1) COVID-19 daily transmission rate per 100K population and (2) COVID-19 mortality rates per 100K population. For the transmission rate analysis, we tested two alternative functional forms: daily cases per 100 thousand people and 7-day moving average of cases per 100 thousand people. The moving average data is likely to be less volatile and serves as a stability test for the daily cases model. The reader would note that we used a natural logarithmic transformation for all the dependent variables. The COVID-19 dataset from Center for Systems Science and Engineering (CSSE) Coronavirus Resource Center at Johns Hopkins University^21^ provides information on the daily confirmed COVID-19 cases, number of people recovered (when available) and the number of deaths from COVID-19 starting from January 22^nd^ to the current date across 3,142 counties in the United States. In our research, we confined our analysis to the cases between March 25^th^ to July 3^rd^ resulting in 101 days of data. Further, we focus on counties that have at least 100 cases by July 3^rd^ and have available information on the mobility trends. With this requirement, a total of 1,258 counties are included in the analysis providing a coverage of 87% of the total population in the United States. For mortality rate, we considered the fatalities within the same time frame across all the 1,258 counties as the transmission rate variable. The summary statistics of the dependent variable are presented in bottom row panel of Table 1.

##### Data Analysis

For the analysis of daily COVID-19 transmission rate, we have repeated measures of the variable (101 repetitions for each county). The traditional linear regression model is not appropriate to study data with multiple repeated observations^22^. Hence, we employ a linear mixed modeling approach that builds on the linear regression model while incorporating the influence of repeated observations from the same county. A brief description of the linear mixed model is provided in the Supplementary Material. For modeling the COVID 19 mortality rate, we rely on simple linear regression approach as the dependent variable here is the total number of COVID-19 deaths per 100K population at a county level. Both models are estimated in SPSS.

## RESULTS

### COVID-19 Transmission Rate Model Results

The estimation results for the linear mixed model are presented in Table 2^2^.

**Table 2.** Estimation Results for Daily COVID-19 Transmission Rate per 100K Population.

| **Variables** | **Estimates** | **t-statistic** | **p-value** |
| --- | --- | --- | --- |
| Constant | -6.071 | -16.215 | 0.000 |
| **Demographics** |  |  |  |
| % of Female population | 0.024 | 7.496 | 0.000 |
| % Young population (<=18 years) | 0.005 | 2.524 | 0.012 |
| % of Black population | 0.007 | 15.061 | 0.000 |
| % of People less than high school education | 0.025 | 18.454 | 0.000 |
| Ln (median income) | 0.396 | 12.650 | 0.000 |
| Employment rate per capita | 1.409 | 9.640 | 0.000 |
| Ln (% of People living in rural areas) | -0.309 | -9.249 | 0.000 |
| **Health Indicators** |  |  |  |
| Ln (HIV rate per 100K People) | 0.052 | 6.306 | 0.000 |
| Hepatitis C rate per 100K People | 0.027 | 5.152 | 0.000 |
| **Mobility Trends** |  |  |  |
| Ln (Daily Average Exposure), 10 days lag |  |  |  |
| From April 25th | 0.027 | 9.618 | 0.000 |
| % People staying at home |  |  |  |
| 14 days lag | -1.027 | -7.690 | 0.000 |
| **Health Care Infrastructure Attributes** |  |  |  |
| Hospital per 100K People | 0.028 | 8.958 | 0.000 |
| Ln (Testing), 5 days lag |  |  |  |
| March 25<sup>th</sup> to May 10th | 0.020 | 10.481 | 0.000 |
| After May 10th | 0.016 | 7.155 | 0.000 |
| **Temporal Factors** |  |  |  |
| Temporal Lagged Variables |  |  |  |
| 7 days lag (March 25<sup>th</sup> to June 21<sup>st</sup>) | 0.205 | 68.500 | 0.000 |
| 7 days lag (After June 21<sup>st</sup>) | 0.340 | 60.026 | 0.000 |
| 14 days lag | 0.148 | 50.225 | 0.000 |
| Day is Weekend | -0.033 | -5.925 | 0.000 |
| **Correlation** |  |  |  |
| $\sigma$ | 0.973 | 194.881 | 0.000 |
| $\rho$ | 0.960 | 305.240 | 0.000 |
| $\Phi$ | 0.321 | 83.977 | 0.000 |

#### Socio-demographics

In terms of female population, we find that higher proportion of females in the population has a positive impact on transmission rated. The result is in contrast to earlier studies that show women are less likely to be affected by COVID-19 transmission relative to men^16^. Among age and racial distribution proportions, we found that increased percentage of younger individuals (<18 years) and African-Americans is associated with more transmission(see earlier work for similar findings^11,18^). It has been suggested that African-Americans in general reside in densely populated low income neighborhoods with lower access to amenities and are employed in industries that requires more public exposure^17^. Educational attainment in a county also plays an important role in influencing the COVID-19 transmission. The counties with higher share of individuals with less than high school education are likely to report increased incidence of COVID-In terms of income, we find that higher median income counties have a higher incidence of COVID-19. The effect of income might appear counter-intuitive at first glance. However, it is possible that higher income individuals are more likely to get tested (even in the absence of symptoms) due to higher health insurance affordability. With respect to employment rate, counties with higher employment rate reflect more exposure and have a positive association with transmission. The percentage of people living in rural area offers a negative association with the daily COVID-19 incidence. This is intuitive as rural areas are sparsely populated and hence have more opportunity for social distancing thus lowering transmission rates.

#### Health indicators

With respect to health indicators, we tried several variables in the transmission rate model. Of these, two variables - number of people suffering from HIV and hepatitis C in a county offered significant impacts. We observe that counties with higher percentage of HIV and hepatitis C patients have an increased incidence of COVID-19 transmission. Individuals with these diseases have weaker immune systems and hence are more susceptible to COVID-19 transmission.

#### Mobility Trends

In terms of mobility trends, we tested two measures: daily average exposure and percentage of people staying at home. In considering these variables in the model, we recognize that exposure will have a lagged effect on transmission i.e. exposure to virus today is likely to manifest as a case in the next 5 to 14 days. In our analysis, we tested several lag combinations and selected the 10 day lag exposure as it offered the best fit. The exposure variable offers interesting results. Until April 25^th^, exposure variable does not have any impact on transmission. This trend strongly coincides with the lower exposure trends (see Figure 2). After April 25^th^, increased exposure is associated with higher transmission rates 10 days into the future (see Hamada and colleagues^19^ for similar findings). For the second measure, staying at home with 14 days lag, we find that daily transmission rates are affected as expected^4,10^. The reader would note that the two measures considered were not found to be correlated and thus were simultaneously considered in the model.

#### Health Care Infrastructure Attributes

From Table 2, we find that counties with more hospitals per capita are more likely to report higher COVID-19 transmission rate. This result perhaps accounts for the higher availability of COVID-19 testing. The last set of variables within this category corresponds to COVID-19 testing effects. Again, we select a 5 day lag as testing results are likely to be reported in 3-5 days. The coefficient of this variable is positive as expected and highly significant^4^. However, after May10^th^, the effect has a lower magnitude, which suggests that compared to the previous time period (before May 10^th^), higher testing rate will increase the daily COVID-19 transmission at a marginally lower rate.

#### Temporal factors

With data available for 101 days, we can evaluate the effect of the transmission rate in previous days on the current day. As expected, we find a positive association between the daily COVID-19 transmission rate and the number of cases 7 and 14 days prior. The result suggests higher transmission rate in previous time periods (7 and 14 days earlier) is likely to result in increased transmission. However, the effect is higher for the 7 day lagged variable, as evidenced by the higher magnitude associated with the corresponding time period in Table 2. Further, the 7 day lagged transmission rate after June 21^st^ implies a higher impact perhaps explaining the sudden surge in COVID-19 cases in recent weeks. Finally, the weekend variable highlights that the COVID-19 transmission rate is lower during weekends possibly because of reduced testing rate on weekends^23^.

#### Correlation

As indicated earlier, we developed the mixed linear model for estimating the daily COVID-19 transmission rate per 100,000 people while incorporating the dependencies across each county for various repetition levels (such as week and month). We found that the model accommodating weekly correlations provided the best result in terms of statistical data fit and variable interpretation. The final set of variables in Table 2 corresponds to the correlation parameter across every 7 days within a county.

##### COVID-19 Mortality Rate

The coefficients in Table 3 represent the effect of different independent variables on COVID-19 mortality rate at a county level.

**Table 3.** Estimation Results for COVID-19 Mortality Rate per 100K Population.

| **Variables** | **Estimates** | **t-statistic** | **p-value** |
| --- | --- | --- | --- |
| Constant | -6.952 | -7.715 | 0.000 |
| **Demographics** |  |  |  |
| Older people % (>65 years old) | 0.068 | 6.649 | 0.000 |
| Black people% | 0.018 | 5.749 | 0.000 |
| % of People less than high school education | 0.050 | 6.153 | 0.000 |
| Employment rate per capita | 8.369 | 7.853 | 0.000 |
| Income inequality ratio | 0.274 | 4.795 | 0.000 |
| Ln (% of People living in rural areas) | -0.772 | -3.173 | 0.002 |
| **Health Indicators** |  |  |  |
| Ln (HIV rate per 100K people) | 0.082 | 1.557 | 0.120 |
| Cancer rate per 100K people | 0.003 | 3.397 | 0.001 |
| % People having Asthma | 0.091 | 2.720 | 0.007 |
| Cardiovascular disease per 1K people | 0.007 | 2.304 | 0.021 |
| **Health Care Infrastructure Attributes** |  |  |  |
| ICU beds per capita | -0.009 | -4.794 | 0.000 |

#### Socio-demographics

A higher percentage of older people in a county leads to an increased COVID-19 mortality rate (see ^14,18^ for similar findings). Further, consistent with previous research^17^, the current analysis also found that the percentage of African-Americans is positively associated with COVID-19 mortality rate. The variable specific to education attainment indicates that the likelihood of COVID-19 mortality increases with increasing share of people with less than high school education in a county. We find that counties with higher income inequality are more likely to experience higher number of COVID-19 deaths per capita relative to the counties with lower income disparities^24^. Finally, higher employment rate has a positive association with COVID-19 mortality rate.

#### Health Indicators

Several variables significantly influence the COVID-19 mortality rate in a county. For instance, in comparison to other counties, counties with higher number of HIV, cancer, asthma and cardiovascular patients are more likely to have higher number of COVID-19 deaths. This is expected^25–28^ as people with such conditions usually have weaker immune system which makes them vulnerable to the disease.

#### Health Care Infrastructure Attributes

The number of ICU beds per capita at a county is found to have a negative impact on COVID-19 mortality rate suggesting a reduced death rate with higher number of ICU bed per person in a county. The result is intuitive as more ICU bed per capita indicates the county is well equipped to handle higher patient demand and treatment is accessible to more COVID-19 patients.

##### Policy Implications

To illustrate the applicability of the proposed COVD-19 transmission model, we conduct a scenario analysis exercise by imposing mobility restrictions. While earlier researchers explored the influence of mobility measures, these models did not account for county level factors such as socio-demographics, health indicators and hospital infrastructure attributes. In our framework, the sensitivity analysis is conducted while controlling for these factors. The hypothetical restrictions on mobility are considered through the following changes to two variables:

1. county level average daily exposure reduced by 10%, 25% and 50%
2. county level percentage of stay at home population increased to 40%, 50% and 60%. The changes to the independent variables were used to predict the dependent variable.

Subsequently, the variable was converted to the daily cases per 100 thousand people. The results from this exercise are presented in Table 4. We present the average change in cases for all counties (1,258), and for the 25 counties with the highest overall transmission rates. From Table 4, two important observations can be made. First, changes to average daily exposure and stay at home population influence COVID-19 transmission significantly. In fact, by increasing stay at home population share to 50% the model predicts a reduction of the number of cases by about 30%.

**Table 4.** Policy Scenario Analysis of Social Distancing in COVID-19 Transmission Rate per 100K Population.

| **Hypothetical Scenarios** | **1,258 Counties** | **Worst 25 Counties** |
| --- | --- | --- |
| 1: daily average exposure reduced by 10% | -0.225 | -0.240 |
| 2: daily average exposure reduced by 25% | -0.612 | -0.653 |
| 3: daily average exposure reduced by 50% | -1.465 | -1.565 |
| 4: 40% people stay at home | -22.949 | -23.206 |
| 5: 50% people stay at home | -30.469 | -30.701 |
| 6: 60% people stay at home | -37.256 | -37.465 |

Second, the benefit from mobility restrictions is slightly higher for the 25 counties with higher overall cases. The two observations provide evidence that issuing lockdown orders in counties with a recent surge is a potential mitigation measure to curb future transmission.

The COVID-19 total mortality rate model can be employed to identify vulnerable counties that need to be prioritized for vaccination programs (when available). While prioritizing the counties based on mortality rate might be a potential approach, it might not always be feasible. To elaborate, vaccination programs have to be planned well in advance (say 2 months) of the vaccine availability. As total mortality rates for 2 months into the future are unavailable, we need a model to predict total mortality into the future. The estimated mortality rate model provides a framework for such analysis. To be sure, it would be prudent to update the proposed model with the latest data to develop a more accurate prediction system.

## DISCUSSION

The current study develops a comprehensive framework for examining COVID-19 transmission and fatality rates in the United States at a county level including an exhaustive set of independent variables: socio-demographics, health indicators, mobility trends and health care infrastructure attributes. In our analysis, we consider all counties with total number of cases greater than 100 on July 3^rd^ and analyze daily cases data from March 25^th^ to July 3^rd^, 2020. The COVID-19 transmission rate is modeled at a daily basis using a linear mixed method while the total mortality rate is analyzed adopting a linear regression approach.

Several county level factors including proportion of African-Americans, income inequality, health indicators associated with Asthma, Cancer, HIV and heart disease, percentage of stay at home individuals, testing infrastructure and Intensive Care Unit capacity impact transmission and/or mortality rates. The results clearly support our hypothesis of considering a universal set of factors in analyzing the COVID-19 data. Further we conducted policy scenario analysis to evaluate the influence of social distancing on the COVID-19 transmission rate. The results highlight the effectiveness of social distancing in mitigating the virus transmission. In fact, we found that by increasing stay at home population share to 50% the model predicts a reduction of the number of cases by about 30%. The finding provides evidence that issuing lockdown orders in counties with a recent surge is a potential mitigation measure to curb future transmission.

To be sure, the study is not without limitations. The study is focused on county level analysis and is intended to reflect associations as opposed to causation. For such causation based analysis, data from individuals would be more suitable. While exposure data were reasonably addressed, data was not available for mask wearing behavior across all counties. Finally, the data on transmission and mortality are updated for few counties to correct for errors or omissions. These were carefully considered in our data preparation. However, it is possible that further updates might be made after we finished our analysis.

## Data Availability

All the data regarding the COVID cases are collected from Center for Systems Science and Engineering (CSSE) Coronavirus Resource Center at Johns Hopkins University.
For independent varaibles, The socio-demographic variables are collected from the American Community Survey; The health indicator variables are collected from the Centers for Disease Control and Prevention (CDC) systems; information on social distancing is collected from Safegraph data; Information about the hospitals and ICU beds are gathered from the County level health ranking data; COVID-19 testing measures are sourced from the COVID-19 tracking project

https://coronavirus.jhu.edu/map.html

https://covidtracking.com/data/.

https://www.countyhealthrankings.org/

## Contributors

NE conceptualized the study. TB and NE finalized the study design. TB, SD and NC conducted the literature review. TB, SD and NC collected the data. TB, SD, NC, and NE analyzed and interpreted the model results. TB, NC and SD prepared the figures. TB, SD, NC and NE drafted the main manuscript. All authors reviewed the results and approved the final version of the manuscript.

## Acknowledgement

The authors would like to gratefully acknowledge SafeGraph COVID-19 Data Consortium, County Health Ranking and Road Maps, Centers for Disease Control System for providing access to the data at county level for United States.

## Footnotes

1 SafeGraph is a data company that aggregates anonymized location data from numerous applications in order to provide insights about physical places. To enhance privacy, SafeGraph excludes census block group information if fewer than five devices visited an establishment in a month from a given census block group

2 As discussed earlier, we also developed the same mixed linear model to estimate the 7-day moving average of COVID-19 cases per capita and find similar results as in the daily COVID-19 transmission model (results are available upon request from the authors). This further reinforces the stability of the transmission model.

